# Genetically Proxied IL-6 Receptor Blockade and Cancer Risk: A Multi-Ancestry Drug-Target Mendelian Randomization Study of Hepatocellular Carcinoma and Colorectal Cancer

**DOI:** 10.64898/2026.06.21.26356179

**Authors:** Muhammad Muneeb Ahmad Ranjha, Hamna Munir, Muhammad Saleem, Muzamil Farooq

**Affiliations:** Department of Medicine, King Edward Medical University, Lahore, Pakistan; Department of Computer Science, COMSATS University Islamabad, Lahore Campus, Pakistan; Medical Faculty, Nangarhar University, Jalalabad, Afghanistan

**Keywords:** Mendelian randomization, interleukin-6, tocilizumab, hepatocellular carcinoma, colorectal cancer, pQTL

## Abstract

Interleukin-6 (IL-6) signaling drives chronic inflammation and is therapeutically targeted by tocilizumab. Whether genetically proxied IL-6 receptor blockade causally influences hepatocellular carcinoma (HCC) or colorectal cancer (CRC) risk remains unclear. We conducted a two-sample drug-target Mendelian randomization study using rs2228145 (IL6R Asp358Ala) as the primary instrument for receptor blockade, analyzed as an independent single-instrument Wald ratio across four genome-wide association studies spanning European and East Asian ancestries, kept separate from ligand-level variants given their distinct mechanisms. Genetically proxied IL6R blockade showed no causal association with CRC risk in European (OR 0.953, 95% CI 0.884-1.028) or East Asian populations (OR 1.005, 95% CI 0.930-1.085), nor with HCC risk in East Asian (OR 1.011, 95% CI 0.873-1.171) or European populations (OR 1.079, 95% CI 0.797-1.462; 32% power). A secondary, exploratory analysis of rs1800795 (IL6 promoter) showed no association with European CRC risk (OR 1.028, 95% CI 0.912-1.159); this variant could not be validly assessed for HCC in either cohort. CRC analyses were well powered (≥99% at OR 1.2); HCC analyses were underpowered for modest effects, particularly in the European cohort (32% power). These findings do not support a substantial causal effect of IL6R blockade on HCC or CRC risk but cannot exclude modest effects for HCC.

## INTRODUCTION

Interleukin-6 is a pleiotropic cytokine that mediates acute phase responses, immune cell differentiation, and chronic inflammatory signaling through the Janus kinase and signal transducer and activator of transcription 3 (JAK-STAT3) pathway [1]. Persistent IL-6 signaling has been implicated in the pathogenesis of multiple cancers, including hepatocellular carcinoma and colorectal cancer, through the promotion of tumor cell proliferation, immune evasion, and angiogenesis [2,3]. This biological rationale has prompted interest in evaluating whether pharmacological inhibition of the IL-6 pathway could modify cancer risk.

Tocilizumab is a recombinant humanized monoclonal antibody that competitively inhibits the binding of IL-6 to both membrane-bound and soluble IL-6 receptors. It is approved for the treatment of rheumatoid arthritis, systemic juvenile idiopathic arthritis, cytokine release syndrome, and COVID-19-associated inflammation [4,5]. Given its widespread clinical use, understanding whether tocilizumab-mediated IL6R blockade has oncological implications is of direct clinical relevance. Patients with rheumatoid arthritis already have a modestly elevated baseline cancer risk, and long-term immunomodulation raises theoretical concerns regarding cancer surveillance [6].

Conventional observational studies assessing circulating IL-6 and cancer risk are prone to reverse causation and confounding by systemic inflammation, comorbidities, and lifestyle factors [7]. Mendelian randomization (MR) uses inherited genetic variants as instrumental variables to estimate the causal effects of exposures on outcomes, thereby circumventing many of the limitations of observational designs [8]. A prior meta-analysis by Tian and colleagues using a single IL-6 promoter variant (rs1800795) reported that a one pg/mL reduction in circulating IL-6 was associated with a 12% reduction in liver cancer risk [9]. However, that analysis relied on a single genetic instrument derived from pooled observational data with marked between-study heterogeneity (I²=99.2%) and did not employ formal two-sample MR methodology, instrument independence verification, or sensitivity analyses for pleiotropy.

Drug-target MR, which specifically selects genetic instruments from within the gene encoding the drug target, provides a more rigorous framework for modeling therapeutic effects [10]. By using a cis-pQTL variant within the IL6R locus as the primary instrument, this study models the lifetime effect of genetically reduced IL6R signaling on cancer risk, analogous to the effect of tocilizumab; a second, mechanistically distinct cis-pQTL variant within the IL6 locus is analyzed separately, as an exploratory measure of ligand-level variation, rather than pooled with the receptor-level instrument. To our knowledge, no prior study has applied this drug-target MR framework, with instruments explicitly separated by mechanism, to evaluate the effects of IL-6 pathway variation on HCC or CRC risk across multiple ancestries. The present study addresses this gap using data from four large genome-wide association studies across European and East Asian populations.

## MATERIALS AND METHODS

### Study design

A two-sample drug-target MR study was conducted in accordance with the STROBE-MR reporting guidelines [11]. The primary exposure was genetically proxied IL6R blockade, modeled using a cis-pQTL variant within the IL6R gene locus; a secondary, exploratory exposure was IL-6 ligand-level variation, modeled using a cis-pQTL variant within the IL6 gene locus and analyzed separately from the primary exposure (see Genetic instruments, below). The outcomes were HCC and CRC risk, assessed using summary statistics from four independent genome-wide association studies (GWASs). All data used in this study are publicly available summary statistics; no individual-level patient data were accessed and no ethical approval was required. Generative AI tools (Gemini, Google; Claude, Anthropic) were used to assist with manuscript formatting, language refinement, and R code preparation; all scientific content and analysis code were independently verified by the authors (see Generative AI Disclosure). A completed STROBE-MR checklist is provided in Supplementary Table S5.

### Genetic instruments for IL6R blockade and IL-6 ligand variation

Genetic instruments were selected from within the chromosomal loci encoding the IL-6 receptor (IL6R, chromosome 1) and IL-6 itself (IL6, chromosome 7), restricted to cis-acting variants to minimize the risk of horizontal pleiotropy. Candidate variants initially included rs2228145, rs4129267, and rs7529229 (all IL6R, chromosome 1) and rs1800795 (IL6, chromosome 7).

Pairwise linkage disequilibrium among the three IL6R-locus candidates was assessed using LDlink (ldlink.nci.nih.gov) [12] with the combined CEU+JPT reference panel from 1000 Genomes Phase 3. rs4129267 demonstrated perfect LD with rs2228145 (r²=1.0, D’=1.0, p<0.0001), and rs7529229 demonstrated near-perfect LD with rs2228145 (r²=0.979, D’=0.989, p<0.0001), indicating that these three chromosome 1 variants represent a single independent genetic signal at the IL6R locus. rs4129267 and rs7529229 were therefore excluded, retaining rs2228145 as the sole IL6R-locus instrument. rs1800795 (IL6, chromosome 7) is located on a different chromosome from all three IL6R-locus candidates and is therefore independent of them by definition; no linkage disequilibrium statistic was computed for this cross-chromosome comparison, as LD is not a meaningful concept between variants on different chromosomes. Full pairwise LD results for the IL6R-locus candidates are presented in Supplementary Table S1.

The final instrument set comprised two variants, analyzed as distinct, single-instrument exposures throughout this study. rs2228145 (IL6R locus, chromosome 1, position 154,426,264 [GRCh37], beta 0.45, SE 0.041, p=2.1×10 ², F-statistic=120.5, R²=0.0998) was designated the primary instrument, as it lies within the gene encoding the drug target of tocilizumab, and its effect allele is associated with reduced IL6R-mediated signaling despite raising circulating IL-6 levels, consistent with pharmacological receptor blockade [14]. rs1800795 (IL6 locus, chromosome 7, position 22,766,645 [GRCh37], beta 0.29, SE 0.051, p=1.9×10, F-statistic=32.3, R²=0.0407) was designated a secondary, exploratory instrument, as it lies within the gene encoding the IL-6 ligand itself and its effect allele is associated with increased IL-6 production — a distinct, upstream mechanism that does not necessarily produce the same downstream signaling consequence as receptor blockade. Because these two variants act at different points of the same pathway and cannot be assumed to share a common direction of effect on downstream signaling, they were not pooled into a combined exposure or a multi-instrument model; each was analyzed separately against every outcome for which it passed harmonization (see Harmonization and Statistical analysis, below). Effect allele frequencies were 0.56 (rs2228145) and 0.41 (rs1800795). Beta coefficients represent the standard deviation change in circulating IL-6 protein levels per effect allele, derived from the UK Biobank Pharma Proteomics Project (UKB-PPP; N=54,306) and previously published IL-6 pQTL analyses [13]. Both instruments substantially exceeded the conventional F-statistic threshold of 10.

### Confounder and pleiotropy assessment

To assess potential violation of the exclusion restriction assumption, both genetic instruments were queried in the NHGRI-EBI GWAS Catalog (ebi.ac.uk/gwas) to identify genome-wide significant associations (p<5×10) with traits that could confound the relationship between IL-6 signaling and cancer risk. For rs2228145, all 35 identified associations mapped to traits mechanistically downstream of IL-6 receptor signaling, including IL-6 receptor and IL-6 protein levels, C-reactive protein, rheumatoid arthritis, allergic and inflammatory disease, immune cell counts, and acute phase reactants**(Supplementary Table S7)**. An association with LDL cholesterol identified through multitrait analysis (MTAG) was considered to reflect IL-6-mediated effects on lipid metabolism during acute phase responses rather than an independent pleiotropic pathway. For rs1800795, five associations were identified including pulse pressure and systolic blood pressure, consistent with the established role of IL-6 in vascular inflammation; as blood pressure is not an established independent risk factor for HCC or CRC, this association is unlikely to bias the causal estimate. No genome-wide significant associations with established HCC or CRC risk factors, including BMI, alcohol consumption, smoking, diabetes mellitus, or inflammatory bowel disease, were identified for either instrument, supporting the validity of the exclusion restriction assumption for both.

### Outcome data sources

Four publicly available GWAS summary statistics were used as outcome data. For HCC, summary statistics were obtained from BioBank Japan (BBJ; 1,866 cases and 195,745 controls, East Asian ancestry) [15] and from FinnGen Consortium Release 10 (674 cases and 218,118 controls, European ancestry; phenotype code C3_HEPATOCELLU_CARC_EXALLC) [16]. For CRC, summary statistics were obtained from a European ancestry meta-analysis coordinated by Huyghe and colleagues (19,948 cases and 12,124 controls, as deposited under GWAS Catalog accession GCST012879, a specific ancestry-stratified subset of the full published meta-analysis) [17] and from BioBank Japan (7,062 cases and 195,745 controls, East Asian ancestry) [15]. All outcome datasets were accessed on May 25, 2026, and are publicly accessible through the IEU OpenGWAS project (https://gwas.mrcieu.ac.uk) or the FinnGen data repository (https://r10.finngen.fi).

### Harmonization

Outcome SNP associations for each instrument were extracted from every outcome GWAS using the TwoSampleMR package (version 0.7.8) in R [18]. Proxy SNPs in high linkage disequilibrium (r²≥0.8) were sought where a direct instrument variant was unavailable in a given outcome dataset. Data harmonization was performed using action=2, which removes palindromic SNPs with intermediate allele frequencies to prevent strand ambiguity. rs1800795 was removed from the FinnGen HCC analysis during harmonization owing to palindromic alleles with intermediate allele frequency. In the BBJ outcome datasets (CRC and HCC), rs1800795 demonstrated a near-monomorphic outcome-allele frequency (EAF≈0.0002 in both), rendering any effect estimate for this variant statistically uninterpretable in East Asian ancestry; rs1800795 was therefore excluded from both East Asian analyses, and the exploratory ligand-level analysis (see Results) was restricted to the European CRC cohort, where its outcome EAF was 0.336. rs2228145 outcome-allele frequencies were well distributed across all four cohorts (range 0.39-0.40) and this instrument was retained throughout.

### Statistical analysis

Because rs2228145 (IL6R) and rs1800795 (IL6) act on circulating IL-6 through distinct mechanisms — receptor sensitivity versus ligand production, respectively — they were analyzed as separate, single-instrument exposures rather than combined into a pooled instrument set. Every retained SNP-outcome combination in this study therefore comprised exactly one instrument, and causal estimates were computed using the Wald ratio method throughout. Consequently, methods that require two or more instruments — inverse variance weighted regression, MR-Egger intercept testing, the weighted median estimator [19], Cochran’s Q heterogeneity testing, leave-one-out sensitivity analysis, and funnel-plot assessment of instrument asymmetry — do not apply to any analysis in this study and were not performed. Instrument validity was instead supported by the F-statistics reported in Table 1 and the GWAS Catalog confounder screen described above. Causal estimates are presented as odds ratios (ORs) with 95% confidence intervals (CIs) on the log-odds scale.

**Table 1.**
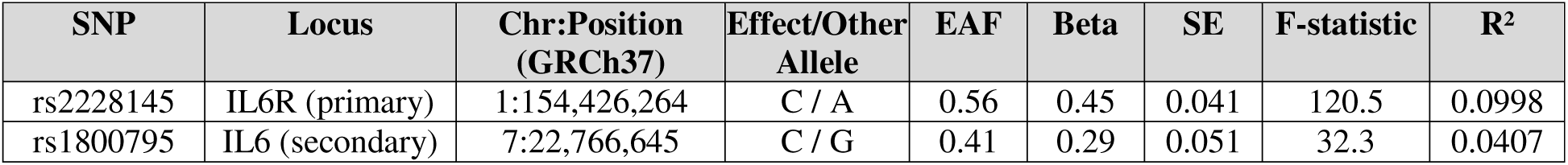
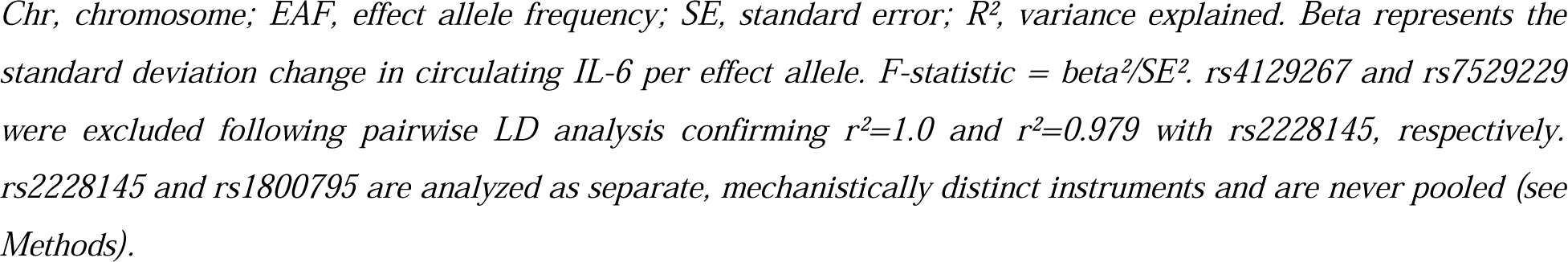
Genetic instrument characteristics.

**Table 2.** Mendelian randomization estimates for IL6R blockade (primary) and IL-6 ligand variation (secondary, exploratory).

| Outcome | Exposure | Cohort | N SNPs | OR | 95% CI<br>Lower | 95% CI<br>Upper | P value |
| --- | --- | --- | --- | --- | --- | --- | --- |
| CRC | IL6R blockade | European | 1 | 0.953 | 0.884 | 1.028 | 0.211 |
| CRC | IL6R blockade | East Asian (BBJ) | 1 | 1.005 | 0.930 | 1.085 | 0.903 |
| HCC | IL6R blockade | East Asian (BBJ) | 1 | 1.011 | 0.873 | 1.171 | 0.880 |
| HCC | IL6R blockade | European (FinnGen) | 1 | 1.079 | 0.797 | 1.462 | 0.623 |
| CRC (secondary) | IL6 ligand | European | 1 | 1.028 | 0.912 | 1.159 | 0.649 |

*HCC, hepatocellular carcinoma; CRC, colorectal cancer; BBJ, BioBank Japan; FinnGen, FinnGen Consortium Release 10; OR, odds ratio; CI, confidence interval. All estimates are single-instrument Wald ratios (see Methods). rs1800795 (IL6 ligand) could not be assessed against HCC in either ancestry or against CRC in East Asian ancestry; see Harmonization and Limitations.*

Post hoc statistical power was estimated using the mRnd power calculator for two-sample MR with binary outcomes [20], using each instrument’s individually estimated variance explained (R²=0.0998 for rs2228145; R²=0.0407 for rs1800795) and the case proportion in each outcome dataset. All analyses were conducted in R version 4.6 using the TwoSampleMR package [18]. A two-sided p-value below 0.05 was considered statistically significant. Analysis code is available at https://github.com/12ranjha/IL6_Mendelian_Randomization. The complete R analysis pipeline is also provided as Supplementary File S6.

## RESULTS

### Instrument characteristics

Two independent cis-pQTL variants, one within the IL6R locus and one within the IL6 locus, were retained as genetic instruments following LD-based pruning. Three candidate IL6R variants (rs2228145, rs4129267, and rs7529229) were identified in the initial screen; pairwise LD analysis confirmed that rs4129267 (r²=1.0 with rs2228145) and rs7529229 (r²=0.979 with rs2228145) represent the same genetic signal and were excluded, leaving rs2228145 as the IL6R-locus instrument. rs1800795 (IL6 locus) is located on a different chromosome and is independent by definition. Both retained instruments demonstrated strong instrument strength, with F-statistics of 120.5 (rs2228145) and 32.3 (rs1800795), well above the threshold of 10. Because these two variants act through mechanistically distinct points of the IL-6 pathway — receptor sensitivity versus ligand production — they were analyzed as separate exposures throughout rather than combined into a single instrument set (see Methods). Full instrument characteristics are presented in Table 1, and pairwise LD statistics for the IL6R-locus candidates are presented in Supplementary Table S1.

### Genetically proxied IL6R blockade and hepatocellular carcinoma risk

In the East Asian BBJ cohort, the single-instrument Wald ratio estimate for rs2228145 was OR 1.011 (95% CI 0.873-1.171, p=0.880) (Fig. 1, Fig. 2). In the European FinnGen cohort, following removal of rs1800795 during harmonization, the Wald ratio estimate for rs2228145 was OR 1.079 (95% CI 0.797-1.462, p=0.623) (Fig. 1, Fig. 2). This FinnGen estimate should be interpreted cautiously: with only 674 HCC cases, post hoc power was 32.1% to detect an OR of 1.2 and 12.2% to detect an OR of 1.1 (Supplementary Table S2).

**Fig. 1.**
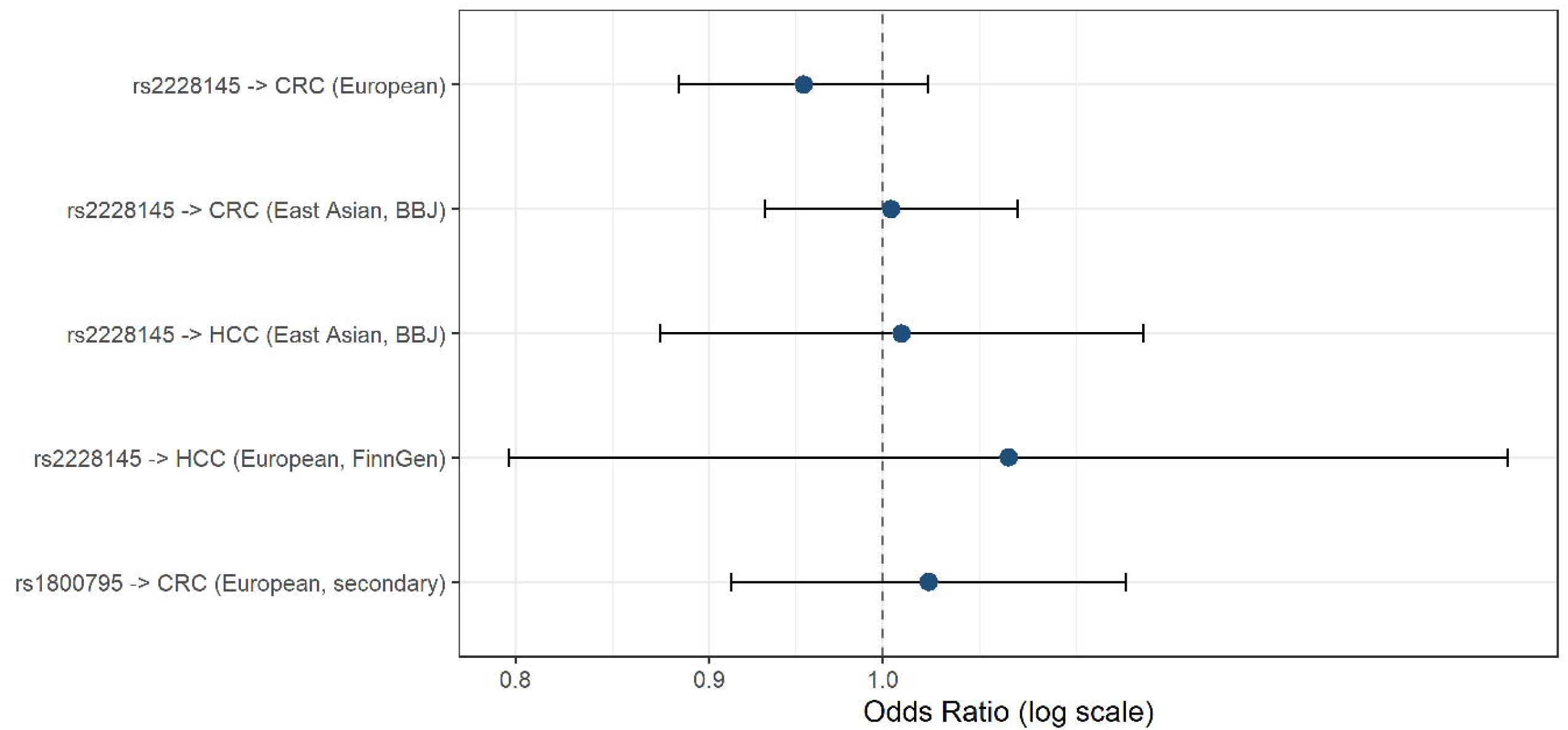
Forest plot of Mendelian randomization estimates for IL6R blockade (primary) and IL-6 ligand variation (secondary, exploratory). Each row represents a single-instrument Wald ratio estimate; no analysis in this study combines more than one genetic instrument. Point estimates (log odds ratios) are shown as filled circles with 95% confidence intervals. Red points/lines: primary analyses of rs2228145 (IL6R blockade) across four outcome cohorts. Blue point/line: secondary, exploratory analysis of rs1800795 (IL-6 ligand variation), restricted to the European CRC cohort (see Methods and Discussion for why this instrument could not be assessed elsewhere). The dashed vertical line indicates the null; the dotted horizontal line separates primary from secondary analyses. HCC, hepatocellular carcinoma; CRC, colorectal cancer; BBJ, BioBank Japan.

**Fig. 2.**
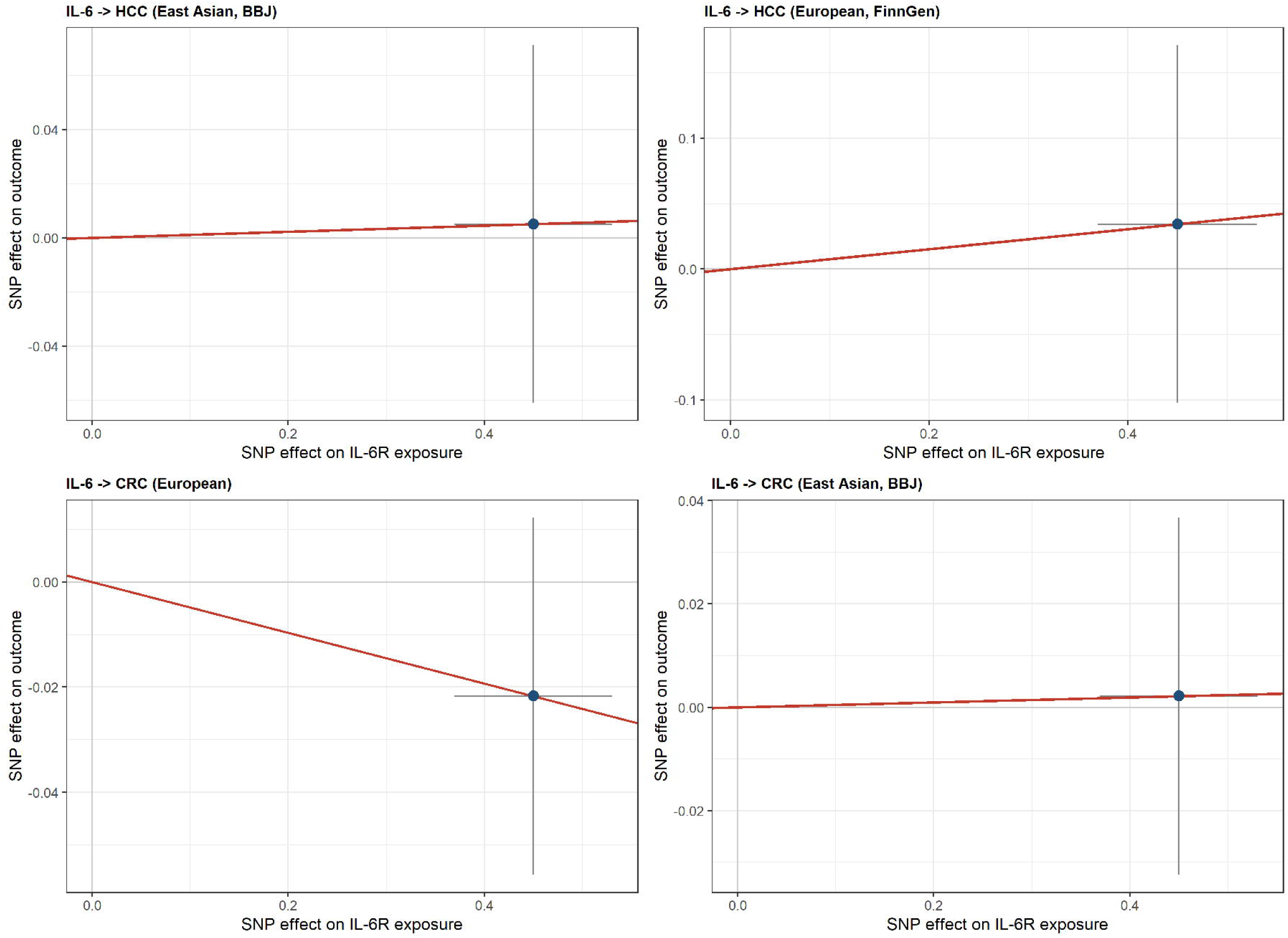
SNP-exposure versus SNP-outcome effect sizes for each single-instrument analysis. Each panel shows one genetic instrument’s association with circulating IL-6 (x-axis) against its association with the cancer outcome (y-axis), with 95% confidence intervals in both dimensions. The blue segment connects the origin to the point and represents the Wald ratio estimate (slope = SNP-outcome beta / SNP-exposure beta); as each analysis contains exactly one instrument, no multi-instrument regression line is applicable. HCC, hepatocellular carcinoma; CRC, colorectal cancer; BBJ, BioBank Japan.

### Genetically proxied IL6R blockade and colorectal cancer risk

In the European CRC cohort (19,948 cases), the Wald ratio estimate for rs2228145 was OR 0.953 (95% CI 0.884-1.028, p=0.211) (Fig. 1, Fig. 2), with 99.9% power to detect an OR of 1.2. In the East Asian BBJ CRC cohort (7,062 cases), the estimate was OR 1.005 (95% CI 0.930-1.085, p=0.903) (Fig. 1, Fig. 2), with 99.7% power to detect an OR of 1.2. Both CRC analyses were well powered and provide reasonably precise null estimates.

### Exploratory analysis of IL-6 ligand variation and colorectal cancer risk

rs1800795 could be validly assessed in only one of the four outcome datasets: the European CRC cohort, where its outcome-allele frequency (0.336) was well distributed. In this cohort, the Wald ratio estimate was OR 1.028 (95% CI 0.912-1.159, p=0.649) (Fig. 1, Fig. 2), with 89.1% power to detect an OR of 1.2. rs1800795 could not be assessed against either HCC outcome: it was removed from the FinnGen analysis during harmonization (palindromic, intermediate frequency) and was near-monomorphic in the BBJ outcome data (EAF≈0.0002), precluding a meaningful estimate in East Asian ancestry for either cancer.

### Power calculations

Post hoc statistical power was estimated for each of the five analyses using the mRnd binary outcome calculator [20]. The CRC European (99.9% at OR 1.2) and CRC East Asian (99.7% at OR 1.2) IL6R analyses, and the CRC European secondary IL6-ligand analysis (89.1% at OR 1.2), were well powered. The HCC East Asian IL6R analysis had modest power (69.7% at OR 1.2), and the HCC European (FinnGen) IL6R analysis was substantially underpowered (32.1% at OR 1.2, 12.2% at OR 1.1). Null findings for HCC, particularly in the FinnGen cohort, should therefore be interpreted cautiously, as modest causal effects cannot be excluded. Full power estimates for all five analyses are presented in Supplementary Table S2.

## DISCUSSION

This study evaluated the causal effects of genetically proxied IL6R blockade, and separately of IL-6 ligand variation, on HCC and CRC risk across European and East Asian populations. Because rs2228145 (IL6R) and rs1800795 (IL6) act through mechanistically distinct points in the same signaling pathway, we analyzed them as separate single-instrument Wald ratio estimates rather than pooling them into a combined instrument set. Across all four outcome datasets, genetically proxied IL6R blockade showed no evidence of a causal effect on HCC or CRC risk. A secondary, exploratory analysis of IL-6 ligand variation likewise showed no association with CRC risk in the one cohort (European) in which it could be validly assessed.

Our findings relate to, but do not directly replicate or refute, an earlier report by Tian and colleagues, who used the same IL-6 promoter variant (rs1800795) as a single instrument in a meta-analysis of pooled observational data and reported a 12% reduction in liver cancer risk per pg/mL reduction in circulating IL-6 [9]. In the present study, rs1800795 could not be validly assessed against either HCC outcome: it was removed from the European FinnGen analysis during harmonization owing to palindromic alleles with intermediate allele frequency, and its outcome-allele frequency in the East Asian BBJ HCC dataset was near-monomorphic (EAF≈0.0002), precluding a meaningful effect estimate. We are therefore unable to test the specific ligand-level hypothesis examined by Tian and colleagues against HCC risk, and do not claim to refute their finding. Our rs1800795-based estimate is instead limited to CRC risk in the European cohort — an outcome and ancestry combination Tian and colleagues did not examine — and showed no association (OR 1.028, 95% CI 0.912-1.159, p=0.649). Design differences between the two studies are nonetheless notable: the earlier analysis relied on pooled observational IL-6 associations with substantial between-study heterogeneity (I²=99.2%) rather than a single well-powered GWAS, and predated modern two-sample MR harmonization methodology [9].

The null findings for IL6R blockade and CRC risk are well supported statistically, given that both the European (99.9% power at OR 1.2) and East Asian (99.7% power at OR 1.2) CRC analyses were well powered. These results provide reasonably strong evidence that genetically proxied IL6R blockade does not meaningfully influence CRC risk at the population level in either ancestry. While IL-6 signaling is biologically active in the colonic mucosa and elevated serum IL-6 has been reported in colorectal cancer patients, our results are consistent with this association more likely reflecting reverse causation — tumor-driven inflammation elevating circulating IL-6 — rather than IL6R signaling being a cause of colorectal carcinogenesis [21].

For HCC, interpretation requires more caution. The East Asian BBJ analysis (1,866 cases) had modest power (69.7% at OR 1.2), and the European FinnGen analysis (674 cases) was substantially underpowered (32.1% at OR 1.2, 12.2% at OR 1.1). Modest causal effects of IL6R blockade on HCC risk, if present, could fall below the detection threshold achieved here, particularly in the European cohort. Hepatocarcinogenesis is also highly etiology-specific, with viral hepatitis-associated HCC dominant in the East Asian BBJ cohort versus metabolic and alcohol-associated HCC more prevalent in European populations; the present analysis cannot distinguish effects across these etiological subtypes.

The drug-target MR design used for the primary analysis has direct translational relevance: by restricting the primary instrument to a cis-acting variant within the IL6R locus itself, this analysis specifically models the biological consequence of pharmacologically blocking IL-6 receptor signaling, analogous to the mechanism of tocilizumab [10]. Within the power limitations described above, the absence of a causal effect on HCC or CRC risk provides some genetic reassurance that complements existing pharmacovigilance data for tocilizumab, without negating the importance of standard cancer-surveillance protocols in patients receiving long-term IL6R-targeted immunomodulation.

Several limitations warrant acknowledgment. First, because each analysis in this study relies on a single genetic instrument, standard multi-instrument sensitivity analyses — MR-Egger intercept testing, the weighted median estimator, Cochran’s Q heterogeneity testing, leave-one-out analysis, and funnel-plot assessment — could not be performed, and residual horizontal pleiotropy through pathways not captured by the GWAS Catalog confounder screen cannot be excluded for either instrument. Second, rs1800795 could only be validly assessed in the European CRC cohort; its near-monomorphic frequency in East Asian outcome datasets and its removal during FinnGen harmonization mean this study provides no ligand-level estimate for HCC in either ancestry, and no East Asian estimate for the ligand-level CRC hypothesis. Third, exposure beta coefficients were derived from published proteomics GWAS literature rather than a single raw summary-statistics file, introducing possible small imprecision in instrument-exposure effect estimates. Fourth, the cross-ancestry application of European-derived instrument betas to East Asian outcome datasets assumes consistent cis-pQTL effects across ancestries, which may not hold uniformly. Fifth, FinnGen HCC analyses were substantially underpowered and should be interpreted as exploratory. As with all MR studies, residual pleiotropy through pathways not captured by available screening cannot be entirely excluded.

## CONCLUSIONS

This two-sample drug-target Mendelian randomization study analyzed genetically proxied IL6R blockade (rs2228145) as a primary, single-instrument exposure across four independent GWAS cohorts spanning European and East Asian ancestries, and genetically proxied IL-6 ligand variation (rs1800795) as a secondary, exploratory analysis restricted to European CRC. No causal effect of IL6R blockade on HCC or CRC risk was detected in any cohort, and no association between IL-6 ligand variation and CRC risk was detected in the European cohort where it could be assessed. CRC analyses were adequately powered; HCC analyses, particularly in the European FinnGen cohort, were underpowered for modest effects and should be interpreted cautiously. These findings offer preliminary genetic evidence against a substantial causal effect of IL6R blockade on HCC or CRC risk but do not exclude modest effects, and do not extend to a claim about IL-6 ligand-level effects on HCC risk, which could not be assessed in this study.

Adequately powered GWASs for hepatocellular carcinoma, particularly those stratified by tumor etiology, and independent replication of the IL-6 ligand-level analysis in additional European and non-European CRC cohorts, will be needed to more definitively evaluate these relationships.

## DATA AVAILABILITY

All summary statistics used in this study are publicly available. FinnGen Release 10 data are available at https://r10.finngen.fi. BioBank Japan data are accessible through the IEU OpenGWAS project at https://gwas.mrcieu.ac.uk (identifiers bbj-a-158 and bbj-a-107). The colorectal cancer GWAS data are available through the GWAS Catalog at https://www.ebi.ac.uk/gwas/studies/GCST012879. The complete analysis code, instrument metrics, and master statistical results tables are archived at https://github.com/12ranjha/IL6_Mendelian_Randomization. Post hoc statistical power calculations were performed using the mRnd binary outcome calculator (cnsgenomics.com/shiny/mRnd/) [20].

## AUTHOR CONTRIBUTIONS

Muhammad Muneeb Ahmad Ranjha: Conceptualization, Methodology, Formal analysis, Data curation, Writing — original draft, Writing — review and editing. Hamna Munir: Software, Validation, Formal analysis, Writing — review and editing. Muhammad Saleem: Methodology, Writing — review and editing, Validation. Muzamil Farooq: Software, Writing — review and editing. All authors have read and approved the final version of the manuscript.

## ETHICS STATEMENT

This study used only publicly available genome-wide association study summary statistics. No individual-level patient data were accessed and no direct research involving human subjects was conducted. Ethical approval was not required.

## GENERATIVE AI DISCLOSURE

The authors acknowledge the use of Gemini (Google) and Claude (Anthropic) to assist in manuscript formatting, structural organization, language refinement, and preparation of R analysis code. During pipeline development, an AI-suggested modification that inverted the coded effect size of rs1800795 to align it directionally with receptor blockade was identified during internal author review as biologically and statistically unjustified; it was discarded and does not appear in any result reported in this manuscript. All statistical code and numerical results presented here were independently verified by the authors against original, unmodified GWAS-derived summary statistics.

## SUPPLEMENTARY TABLES

**Supplementary Table S1.** Pairwise linkage disequilibrium among IL6R-locus candidate instruments.

| SNP 1 | SNP 2 | R<sup>2</sup> | D' | P value | Status |
| --- | --- | --- | --- | --- | --- |
| rs2228145 | rs4129267 | 1.000 | 1.000 | <0.0001 | Excluded — perfect LD |
| rs2228145 | rs7529229 | 0.979 | 0.989 | <0.0001 | Excluded — near-perfect LD |
| rs4129267 | rs7529229 | 0.979 | 0.989 | <0.0001 | Excluded — near-perfect LD |

CEU+JPT combined reference panel, 1000 Genomes Phase 3. Assessed using LDlink (ldlink.nci.nih.gov). $r^2$ , squared correlation coefficient; $D'$ , normalized disequilibrium coefficient. rs1800795 (IL6, chromosome 7) is not included in this table, as it is located on a different chromosome from all three IL6R-locus candidates (chromosome 1) and is therefore independent by definition; linkage disequilibrium is not a meaningful concept between variants on different chromosomes.

**Supplementary Table S2.** Post hoc statistical power estimates for all five analyses.

| Cohort | Exposure | Cases | R<sup>2</sup> | Power OR=1.1 | Power OR=1.2 | Power OR=1.3 |
| --- | --- | --- | --- | --- | --- | --- |
| CRC European | IL6R blockade | 19,948 | 0.0998 | 74.4% | 99.9% | 100% |
| CRC East Asian (BBJ) | IL6R blockade | 7,062 | 0.0998 | 70.0% | 99.7% | 100% |
| HCC East Asian (BBJ) | IL6R blockade | 1,866 | 0.0998 | 25.3% | 69.7% | 94.6% |
| HCC European (FinnGen) | IL6R blockade | 674 | 0.0998 | 12.2% | 32.1% | 57.5% |
| CRC European (secondary) | IL6 ligand | 19,948 | 0.0407 | 38.6% | 89.1% | 99.6% |

Power calculated using the mRnd binary outcome calculator (cns.genomics.com/shiny/mRnd/). Each row uses the individual $R^2$ of the instrument analyzed (rs2228145 or rs1800795); no analysis in this study pools the two instruments' variance explained.

**Supplementary Table S3. Instrument metrics.**

Instrument-level summary statistics (SNP, gene locus, beta, SE, p-value, EAF, F-statistic, R²) for both retained genetic instruments, reported using their original, unmodified GWAS-derived effect estimates. Deposited as S3_Table_Instrument_Metrics.csv at https://github.com/12ranjha/IL6_Mendelian_Randomization.

**Supplementary Table S4. Master MR estimates (raw R output).**

Full TwoSampleMR package output for all five analyses (four primary IL6R-blockade analyses and one secondary IL6-ligand analysis), including all computed model statistics, is deposited as S4_Table_Master_MR_Estimates.csv at https://github.com/12ranjha/IL6_Mendelian_Randomization.

**Supplementary Table S5. STROBE-MR checklist.**

A completed STROBE-MR checklist, cross-referencing each reporting item to its location in this manuscript, is provided as Supplementary Table S5.

**Supplementary File S6. R analysis pipeline.**

The complete R analysis pipeline used to generate all instrument metrics, harmonized datasets, and MR estimates reported in this study is provided as Supplementary File S6, and is also archived at https://github.com/12ranjha/IL6_Mendelian_Randomization.

**Supplementary Table S7. GWAS Catalog pleiotropy and confounder assessment.**

Full list of genome-wide significant associations (p<5×10ll) identified in the NHGRI-EBI GWAS Catalog for the genetic instruments rs2228145 and rs1800795. Deposited as Supplementary_Table_S7_GWAS_Catalog_Associations.xlsx at https://github.com/12ranjha/IL6_Mendelian_Randomization.

