## Supplementary Table S5 for "Genetically Proxied IL-6 Receptor Blockade and Cancer Risk: A Multi-Ancestry Drug-Target Mendelian Randomization Study of Hepatocellular Carcinoma and Colorectal Cancer"

**STROBE-MR Reporting Guidelines Checklist**

| **Item No.** | **Checklist Item** | **Section in Manuscript** |
| --- | --- | --- |
| **1** | **Title and abstract** |  |
| **1a** | **Indicate the study's design with a commonly used term in the title or the abstract.** | **Title; Abstract** |
| **1b** | **Provide an informative and balanced summary of what was done and what was found.** | **Abstract** |
| **2** | **Introduction** |  |
| **2a** | **Explain the scientific background and rationale for the investigation being reported.** | **Introduction (Paragraphs 1–3)** |
| **2b** | **State specific objectives, including any prespecified hypotheses.** | **Introduction (Paragraph 4)** |
| **3** | **Methods** |  |
| **3a** | **Present key elements of study design early in the paper.** | **Materials and Methods: Study design** |
| **3b** | **Describe the setting, locations, and relevant dates, including periods of recruitment, exposure, follow-up, and data collection.** | **Materials and Methods: Outcome data sources** |
| **3c** | **Describe the data sources used and their characteristics.** | **Materials and Methods: Outcome data sources** |
| **3d** | **Explain how the genetic variants were chosen as instrumental variables.** | **Materials and Methods: Genetic instruments for IL6R blockade and IL-6 ligand variation** |
| **3e** | **Describe any steps taken to ensure that the genetic variants satisfy the instrumental variable assumptions.** | **Materials and Methods: Confounder and pleiotropy assessment** |
| **3f** | **Describe all statistical methods, including those used to control for confounding.** | **Materials and Methods: Statistical analysis** |
| **3g** | **Describe any methods used to examine subgroups and interactions.** | ***Not applicable* (No subgroup or interaction analyses were performed)** |
| **3h** | **Explain how missing data were addressed.** | **Materials and Methods: Harmonization** |
| **3i** | **Describe any sensitivity analyses.** | **Materials and Methods: Statistical analysis (*Explicitly noted as not applicable due to single-instrument design*)** |
| **4** | **Results** |  |
| **4a** | **Report the numbers of individuals at each stage of the study.** | **Materials and Methods: Outcome data sources** |
| **4b** | **Give characteristics of study participants and information on exposures and potential confounders.** | **Results: Instrument characteristics; Table 1** |
| **4c** | **Report the associations between the genetic variants and the exposure, and between the genetic variants and the outcome.** | **Table 1; Results (all subsections)** |
| **4d** | **Report the main causal estimates and their precision (e.g., 95% confidence intervals).** | **Table 2; Results (all subsections); Figure 1** |
| **4e** | **Report results from sensitivity analyses and assessments of pleiotropy.** | ***Not applicable* (Single-instrument design precludes multi-instrument sensitivity tests)** |
| **5** | **Discussion** |  |
| **5a** | **Summarize key results with reference to study objectives.** | **Discussion (Paragraph 1)** |
| **5b** | **Discuss limitations of the study, taking into account sources of potential bias or imprecision.** | **Discussion (Paragraph 6)** |
| **5c** | **Give a cautious overall interpretation of results considering objectives, limitations, multiplicity of analyses, results from similar studies, and other relevant evidence.** | **Discussion; Conclusions** |
| **5d** | **Discuss the generalisability (external validity) of the study results.** | **Conclusions** |
| **6** | **Other Information** |  |
| **6a** | **Give the source of funding and the role of the funders for the present study and, if applicable, for the original study on which the present article is based.** | **Funding** |
