## Supplementary File S6 for "Genetically Proxied IL-6 Receptor Blockade and Cancer Risk: A Multi-Ancestry Drug-Target Mendelian Randomization Study of Hepatocellular Carcinoma and Colorectal Cancer"

################################################################################

\# Project: Genetically Proxied Interleukin-6 Inhibition and Cancer Risk

\# Pipeline Script: il6_mr_pipeline_corrected.R

\#

\# Final design (5 analyses total, all single-instrument Wald ratio):

\#	1. rs2228145 -> CRC, European

\#	2. rs2228145 -> CRC, East Asian (BBJ)

\#	3. rs2228145 -> HCC, East Asian (BBJ)

\#	4. rs2228145 -> HCC, European (FinnGen)

\#	5. rs1800795 -> CRC, European (secondary/exploratory)

################################################################################

\# ── STEP 1: INITIALIZE ENVIRONMENT & AUTHENTICATION ───────────────────────────

library(TwoSampleMR)

library(ieugwasr)

library(ggplot2)

library(gridExtra)

library(data.table)

library(dplyr)

output_dir <- "./Genuine_MR_Data_Tables"

if (!dir.exists(output_dir)) dir.create(output_dir)

\# ── STEP 2: CONSTRUCT CIS-PQTL EXPOSURE DATA FRAME ────────────────────────────

il6_exposure_raw <- data.frame(

SNP	= c("rs2228145", "rs1800795"),

beta	= c(0.45, 0.29),

se	= c(0.041, 0.051),

pval	= c(2.1e-28, 1.9e-09),

eaf	= c(0.56, 0.41),

effect_allele = c("C", "C"),

other_allele = c("A", "G"),

chr	= c(1, 7),

pos	= c(154426264, 22766645),

Phenotype	= c("IL6R blockade (rs2228145)", "IL6 ligand (rs1800795)"),

samplesize	= rep(54306, 2),

id.exposure	= c("UKB-PPP_IL6R", "UKB-PPP_IL6"),

stringsAsFactors = FALSE

)

exposure_metrics <- il6_exposure_raw %>%

mutate(

F_statistic = (beta^2) / (se^2),

R2 = (2 * (beta^2) * eaf * (1 - eaf))

)

write.csv(exposure_metrics, file = file.path(output_dir, "S3_Table_Instruments_Metrics.csv"), row.names = FALSE)

exposure_il6 <- TwoSampleMR::format_data(

dat	= il6_exposure_raw,

type	= "exposure",

snp_col	= "SNP",

beta_col	= "beta",

se_col	= "se",

pval_col	= "pval",

eaf_col	= "eaf",

effect_allele_col = "effect_allele",

other_allele_col = "other_allele",

phenotype_col	= "Phenotype",

samplesize_col	= "samplesize",

chr_col	= "chr",

pos_col	= "pos"

)

\# ── STEP 3: EXTRACT & HARMONIZE MULTI-ANCESTRY OUTCOMES ────────────────────────

out_crc_eur <- extract_outcome_data(exposure_il6$SNP, "ebi-a-GCST012879", proxies = TRUE, rsq = 0.8)

out_crc_asn <- extract_outcome_data(exposure_il6$SNP, "bbj-a-107", proxies = TRUE, rsq = 0.8)

out_hcc_asn <- extract_outcome_data(exposure_il6$SNP, "bbj-a-158", proxies = TRUE, rsq = 0.8)

\# CORRECTION: explicitly drop rs1800795 from both BBJ outcome sets here, at the

\# source, rather than relying on harmonise_data() to filter it. This is the

\# fix for Problem 2 -- previously it survived harmonization with mr_keep=TRUE

\# despite EAF ~ 0.0002 in BBJ.

out_crc_asn <- out_crc_asn[out_crc_asn$SNP != "rs1800795", ]

out_hcc_asn <- out_hcc_asn[out_hcc_asn$SNP != "rs1800795", ]

finngen_path <- "finngen_R10_HCC.gz"

if (!file.exists(finngen_path)) {

message("FinnGen file not found in current directory. Please select it from the dialog window...")

finngen_path <- file.choose()

}

if (file.exists(finngen_path)) {

fg_raw <- fread(finngen_path)

fg_snps <- fg_raw[rsids %in% exposure_il6$SNP]

fg_snps$rsid_clean <- sapply(strsplit(fg_snps$rsids, ","), `[`, 1)

out_hcc_eur <- TwoSampleMR::format_data(

as.data.frame(fg_snps), type = "outcome",

snp_col = "rsid_clean", beta_col = "beta", se_col = "sebeta",

pval_col = "pval", effect_allele_col = "alt", other_allele_col = "ref", eaf_col = "af_alt"

)

out_hcc_eur$outcome <- "HCC_FinnGen_European"

\# CORRECTION: HCC-FinnGen is only ever analyzed with rs2228145 per the

\# manuscript design (rs1800795 secondary analysis is CRC-European only).

\# Restrict explicitly rather than relying on whatever happens to be in the file.

out_hcc_eur <- out_hcc_eur[out_hcc_eur$SNP == "rs2228145", ]

} else {

stop("Critical Error: File selection was cancelled or failed.")

}

harm_crc_eur <- harmonise_data(exposure_il6, out_crc_eur, action = 2) # keeps BOTH SNPs (analyses 1 + 5)

harm_crc_asn <- harmonise_data(exposure_il6, out_crc_asn, action = 2) # rs2228145 only (analysis 2)

harm_hcc_asn <- harmonise_data(exposure_il6, out_hcc_asn, action = 2) # rs2228145 only (analysis 3)

harm_hcc_eur <- harmonise_data(exposure_il6, out_hcc_eur, action = 2) # rs2228145 only (analysis 4)

\# Belt-and-suspenders: re-assert the exact instrument sets even after

\# harmonization, in case a proxy SNP for rs1800795 slipped in via proxies=TRUE.

harm_crc_asn <- harm_crc_asn[harm_crc_asn$SNP == "rs2228145", ]

harm_hcc_asn <- harm_hcc_asn[harm_hcc_asn$SNP == "rs2228145", ]

harm_hcc_eur <- harm_hcc_eur[harm_hcc_eur$SNP == "rs2228145", ]

\# CORRECTION: write clean, human-readable outcome labels back onto the

\# harmonized data BEFORE any plotting, so scatter-plot y-axes and any legend

\# text show "Colorectal Cancer (European)" etc., never a raw GWAS ID string.

harm_crc_eur$outcome <- "Colorectal Cancer (European)"

harm_crc_asn$outcome <- "Colorectal Cancer (East Asian, BBJ)"

harm_hcc_asn$outcome <- "Hepatocellular Carcinoma (East Asian, BBJ)"

harm_hcc_eur$outcome <- "Hepatocellular Carcinoma (European, FinnGen)"

\# ── STEP 4: ANALYTICAL CORE — WALD RATIO ONLY, EVERYWHERE ─────────────────────

\# CORRECTION: method_list is hard-locked to "mr_wald_ratio". This is the fix

\# for Problem 1. Even in harm_crc_eur, which has 2 SNPs, mr_wald_ratio computes

\# an independent per-SNP ratio for each row -- it will NEVER pool them into an

\# IVW estimate. That is exactly the two-separate-Wald-ratios design the

\# manuscript describes for the CRC-European arm.

cohort_pipeline <- function(harm_data, label, file_prefix) {

write.csv(harm_data, file = file.path(output_dir, paste0("Raw_Harmonized_Data_", file_prefix, ".csv")), row.names = FALSE)

res <- mr(harm_data, method_list = c("mr_wald_ratio"))

res$OR <- exp(res$b)

res$OR_lower <- exp(res$b - 1.96 * res$se)

res$OR_upper <- exp(res$b + 1.96 * res$se)

res$cohort <- label

\# Attach SNP identity explicitly (mr() output doesn't always retain it in a

\# convenient column name across TwoSampleMR versions), needed for the forest

\# plot labels and for distinguishing the CRC-European primary vs secondary row.

res$SNP <- harm_data$SNP[match(paste(res$id.exposure, res$id.outcome), paste(harm_data$id.exposure, harm_data$id.outcome))]

return(res)

}

c1 <- cohort_pipeline(harm_crc_eur, "Colorectal Cancer (European)", "CRC_EUR")

c2 <- cohort_pipeline(harm_crc_asn, "Colorectal Cancer (East Asian, BBJ)", "CRC_ASN")

c3 <- cohort_pipeline(harm_hcc_asn, "Hepatocellular Carcinoma (East Asian, BBJ)", "HCC_ASN")

c4 <- cohort_pipeline(harm_hcc_eur, "Hepatocellular Carcinoma (European, FinnGen)", "HCC_EUR")

all_results <- bind_rows(c1, c2, c3, c4)

write.csv(all_results, file = file.path(output_dir, "S4_Table_Master_MR_Estimates.csv"), row.names = FALSE)

\# Heterogeneity/pleiotropy are not applicable to single-instrument Wald ratios.

\# We record that explicitly rather than generating placeholder numeric fields.

het_plei_note <- data.frame(

note = "Not applicable: all analyses are single-instrument Wald ratios. Heterogeneity (Q) and pleiotropy (MR-Egger intercept) statistics require >=2 independent instruments per analysis and were not computed."

)

write.csv(het_plei_note, file = file.path(output_dir, "S5_S6_Heterogeneity_Pleiotropy_NotApplicable.csv"), row.names = FALSE)

\# ── STEP 5: CUSTOM VISUALIZATION SUITE (no IVW rows, no funnel, no LOO) ───────

\# Row order + display labels for the forest plot, matching the 5-analysis

\# design table exactly: 4 primary rs2228145 rows + 1 secondary rs1800795 row.

forest_df <- all_results %>%

mutate(

row_label = case_when(

SNP == "rs2228145" & cohort == "Colorectal Cancer (European)" ~ "rs2228145 -> CRC (European)",

SNP == "rs2228145" & cohort == "Colorectal Cancer (East Asian, BBJ)" ~ "rs2228145 -> CRC (East Asian, BBJ)",

SNP == "rs2228145" & cohort == "Hepatocellular Carcinoma (East Asian, BBJ)" ~ "rs2228145 -> HCC (East Asian, BBJ)",

SNP == "rs2228145" & cohort == "Hepatocellular Carcinoma (European, FinnGen)" ~ "rs2228145 -> HCC (European, FinnGen)",

SNP == "rs1800795" & cohort == "Colorectal Cancer (European)" ~ "rs1800795 -> CRC (European, secondary)",

TRUE ~ paste(SNP, "->", cohort)

),

analysis_order = case_when(

row_label == "rs2228145 -> CRC (European)"	~ 1,

row_label == "rs2228145 -> CRC (East Asian, BBJ)"	~ 2,

row_label == "rs2228145 -> HCC (East Asian, BBJ)"	~ 3,

row_label == "rs2228145 -> HCC (European, FinnGen)"	~ 4,

row_label == "rs1800795 -> CRC (European, secondary)"	~ 5,

TRUE ~ 99

)

) %>%

arrange(analysis_order)

\# Sanity check: this MUST be exactly 5 rows. If it isn't, something upstream

\# (a stray proxy SNP, a duplicate harmonization) has broken the design and the

\# script stops rather than silently plotting a wrong figure.

if (nrow(forest_df) != 5) {

stop("Expected exactly 5 Wald-ratio analyses but found ", nrow(forest_df),

". Inspect all_results before plotting -- do not proceed.")

}

forest_df$row_label <- factor(forest_df$row_label, levels = rev(forest_df$row_label[order(forest_df$analysis_order)]))

fig1_forest <- ggplot(forest_df, aes(x = OR, y = row_label)) +

geom_vline(xintercept = 1, linetype = "dashed", color = "grey40") +

geom_errorbarh(aes(xmin = OR_lower, xmax = OR_upper), height = 0.15) +

geom_point(size = 3, color = "#1f4e79") +

scale_x_log10() +

labs(x = "Odds Ratio (log scale)", y = NULL,

title = "Figure 1. Wald Ratio Estimates: IL-6 Pathway Instruments and Cancer Risk") +

theme_bw(base_size = 12) +

theme(plot.title = element_text(size = 11, face = "bold"))

ggsave(file.path(output_dir, "Fig1_Forest_corrected.png"), fig1_forest, width = 9, height = 4.5, dpi = 300)

\# Scatter plots: ONLY the four primary rs2228145 analyses, one point each

\# (SNP effect on exposure vs SNP effect on outcome), with a single Wald-ratio

\# slope line (slope = beta.outcome / beta.exposure) through the origin.

\# No IVW line, because IVW was never computed -- there is nothing to draw.

make_wald_scatter <- function(harm_data, title_text) {

d <- harm_data[harm_data$SNP == "rs2228145", ]

slope <- d$beta.outcome / d$beta.exposure

ggplot(d, aes(x = beta.exposure, y = beta.outcome)) +

geom_hline(yintercept = 0, color = "grey80") +

geom_vline(xintercept = 0, color = "grey80") +

geom_errorbar(aes(ymin = beta.outcome - 1.96 * se.outcome, ymax = beta.outcome + 1.96 * se.outcome), width = 0, color = "grey50") +

geom_errorbarh(aes(xmin = beta.exposure - 1.96 * se.exposure, xmax = beta.exposure + 1.96 * se.exposure), height = 0, color = "grey50") +

geom_abline(intercept = 0, slope = slope, color = "#c0392b", linewidth = 0.8) +

geom_point(size = 3, color = "#1f4e79") +

labs(x = "SNP effect on IL-6R exposure", y = "SNP effect on outcome",

title = title_text) +

theme_bw(base_size = 11) +

theme(plot.title = element_text(size = 10, face = "bold"))

}

p1 <- make_wald_scatter(harm_hcc_asn, "IL-6 -> HCC (East Asian, BBJ)")

p2 <- make_wald_scatter(harm_hcc_eur, "IL-6 -> HCC (European, FinnGen)")

p3 <- make_wald_scatter(harm_crc_eur, "IL-6 -> CRC (European)")

p4 <- make_wald_scatter(harm_crc_asn, "IL-6 -> CRC (East Asian, BBJ)")

fig2_scatter <- arrangeGrob(p1, p2, p3, p4, ncol = 2,

top = "Figure 2. Wald Ratio Scatter Plots (rs2228145, primary instrument)")

ggsave(file.path(output_dir, "Fig2_Scatter_corrected.png"), fig2_scatter, width = 12, height = 9, dpi = 300)

message("Pipeline completed. Exactly 5 Wald ratio analyses generated. ",

"No IVW, no funnel plot, no leave-one-out plot were run or plotted.")
